# Assessment of Environmental Factors Associated with Antibiotic Resistance Genes (ARGs) in the Yangtze Delta, China

**DOI:** 10.1101/2022.12.05.22283137

**Authors:** Jiazheng Miao, Yikai Ling, Xiaoyuan Chen, Siyuan Wu, Sajid Umar, Shixin Xu, Benjamin D. Anderson

## Abstract

The emergence of antimicrobial resistance (AMR) is an urgent and complex public health challenge worldwide. As a sub-problem of AMR, antibacterial resistance (ABR) is of particular concern due to inadequacy of alternative medication. Earlier studies have shown that ABR is not only impacted by antibiotics, but also affected by the interactions between bacteria and their environments. Therefore, to combat ABR in a specific region, local environmental conditions must be investigated to comprehensively understand which environmental factors might contribute to ABR and propose more tailored solutions. This study surveyed environmental contributors of antibiotic resistance genes (ARGs), the parameter for measuring ABR, in the Yangtze Delta. A high abundance of ARGs was detected, despite low antibiotic and heavy metal concentrations. Phosphorus, chromium, manganese, calcium, and strontium were identified as potential key contributors of ARGs. Suppression of ARGs could be realized through decreasing the concentration of phosphorus in surface water. Group 2A light metals (e.g., magnesium and calcium) could be developed as eco-friendly reagents for controlling antibiotic resistance in the future.

## 1. Introduction

Antimicrobial resistance (AMR) refers to the process by which bacteria, parasites, viruses, and fungi evolve mechanisms that protect them from drugs [1]. Due to the increasing prevalence, health effects, and economic impact of AMR, World Health Organization has characterized AMR as “a complex global public health challenge” (p. XIX) that requires urgent action [1]. Among the concerns about AMR, countermeasures against antibacterial resistance (ABR) are of particular urgency because of severe inadequacy of alternative therapeutic options [1]. In 2019, it is estimated that 4.95 million deaths were associated with ABR, and 1.27 million deaths were attributable to ABR [2]. In a recent study that estimates the economic burden of ABR in China, 27.45% of inpatient bacterial infection were antibiotic resistant, which is estimated to have caused a 1.5% increase in inpatient mortality rates and cost $77 billion in related societal expenses in 2017 [3]. What is worse, bacteria are even capable of spreading ABR at a high rate through exchanging ARGs in horizontal gene transfer (HGT) [4]. ARGs are a group of genes that encode the proteins responsible for resisting antibiotics and are frequently carried on mobile genetic elements (MGEs) like plasmids, transposons and integrons [4]. In HGT, one bacterial cell transfer its MGEs to other bacterial cells, facilitating environmental dispersal of the ARGs [4]. Thus, ARG has also been recognized as a new environmental pollutant that poses a threat to human health [4].

Earlier studies have attempted to suppress ABR by reducing antibiotic use to allow non-resistant bacteria to outcompete resistant bacteria [5-7], but reversibility only occurs at a slow rate. Later studies demonstrate a strong association between ABR and environmental factors [8]. In particular, heavy metals are reported to have a co-selection effect on ARGs [9-11]. Because bacteria resist antibiotics and heavy metals through similar strategies (e.g., reduction in permeability, efflux, sequestration, etc.), heavy metals may also pose a selective pressure on ARGs. Furthermore, antibiotic resistant communities in urban lakes have also been identified to be more competitive than non-resistant communities at higher temperatures, but less competitive at higher concentration of magnesium or salinity [8].

Therefore, to effectively understand and mitigate ABR in a specific region, local environmental conditions must be investigated.

The Yangtze Delta is one of the most economically developed regions in China. It consists of 26 cities including Shanghai City, southern part of Zhejiang Province, and northern part of Jiangsu province [12]. In 2018, 154 million people were reported to permanently resides in this area at a high population density of 720 persons/square kilometre [12]. The hygiene of drinking water source in this area (e.g., Yangcheng Lake in Suzhou and Huangpu River in Shanghai) is of vital importance since the water sources provide drinking water for over 150 million residents. Due to its economic and environmental importance, previous studies have investigated the prevalence of ARGs and patterns of antibiotic use in the area.

In 2020, 64 out of 69 types of antibiotics monitored in the surface water in China can be detected in Shanghai, indicating the widespread of antibiotics in the surface water of Shanghai, with concentrations ranging from a few pg L^-1^ to hundreds ng L-1 [13-15]. While confirming the widespread distribution and persistence of antibiotics, these studies estimate low environmental risks for most types of antibiotics [14, 16] by calculating their hazard quotients[17]. However, the environmental risks of enrofloxacin, amoxicillin and oxytetracyclines to aquatic organisms, represented by the risk quotient method [18, 19], are distinctly higher than those in other countries [14]. As for ARGs, tetracycline resistance genes (*tet*) and sulfonamide resistance genes (*sul*) exist in most waterbodies in Yangtze Delta, and β-lactam resistance genes also exist in some regions [20, 21]. For *sul* and *tet* genes, the detection frequency of *sul*I, *sul*II, *tet*A, *tet*G, *tet*M and *tet*O were significantly higher than *sul*III, which was still significantly higher than *tet*C [21]. It was observed that samples with more *sul* genes tend to contain higher levels of total sulfonamides, and the same relationship can be applied to *tet* genes [20].

While these studies focus on the link between ARGs and antibiotic detection. Limited studies have investigated how ARGs are influenced by environmental factors. In this study, we tested surface water samples for ARGs, antibiotics, and dissolved elements to better understand what environmental factors might contribute to the prevalence of ARGs in main water bodies in the Yangtze Delta.

## 2. Methods

### 2.1 Sampling Sites and Sample Collection

In 2021, surface water samples were collected from sampling sites located along a watershed starting at Yangcheng Lake in Suzhou, running along the Wusong River, passing Dianshan Lake in Shanghai, running along the Huangpu River in Shanghai, and ending at an estuary at the Shanghai coastal area. The watershed connects most of the main water bodies in Suzhou and Shanghai. The sampling area contained four subregions: Dianshan Lake (Qingpu District, Shanghai), Huangpu River (Songjiang District, Shanghai), estuary (Jinshan District, Shanghai), and Kunshan (Suzhou).

Surface water samples were collected using brown HDPE bottles (NINGKE, China) that were dipped into the top 10cm of the water body with the goal of collecting 1000ml of water. Water temperature, salinity, and pH of the collected samples were immediately measured and recorded using a salinity and pH meter (BANTE, China) according to the manufacturer’s instructions. Sample bottles were transported on wet ice within 3 hours to Duke Kunshan University where 650ml of each sample was filtered through a 0.45 µm glass microfiber filter membranes (Titan, China). The filtrate and filter membranes were stored stored at a -80°C freezer.

### 2.2 Measurement of ARGs

Cells gathered on the filter membrane were first released in 10ml of ultrapure water. A TaKaRa MiniBEST Bacteria Genomic DNA Extraction Kit was used to extract 240µl of DNA from 10ml of stored sample, according to the manufacturer’s instructions. Quantitative PCR was used to screen extracts for ARGs using the SYBR Green Real-Time PCR assay (TaKaRa Co. Dalian, China) on a Mic qPCR Cycler (BioMolecular Systems, EI Cajon, CA), according to the manufacturer’s instructions. One sample was randomly selected from each of the four subregions. A total of 26 primers for ARGs were used to screen the four samples. The ARGs include 15 tetracycline resistance genes (*tetA, tetC, tetE, tetK, tetL, tetA/P, tetG, tetM, tetO, tetQ, tetS, tetT, tetW, tetBP*, and *tetX*), 4 β-lactam resistance genes (*bla*_CTX-M_, *bla*_TEM_, *bla*_SHV_, and *bla*_ampC_), 3 sulfonamide resistance genes (*sul*I, *sul*II, and *sul*III), 3 macrolides resistance genes (*ereA, ereB*, and *mphA*), and 1 betalactam resistance gene. Primer sequences and cycling conditions for the quantitative PCR experiments are listed in Table S1. Cultured environmental samples were used as positive controls.

Based on the ARG screening results (Figure S1a and Figure S1b), the three most abundant ARGs were selected for further quantitative PCR experiments: *tetBP, ampr, tetO*. Each gene was commercially synthesized and used as an internal standard to determine absolute abundance.

### 2.3 Measurement of Dissolved Elements Concentrations

After filtration, approximately 150ml of stored filtrate was acidified to pH < 2 by (1+1) nitric acid and preserved at -80 °C freezer. The samples were sent to the Nanjing Institute of Geography & Limnology, Chinese Academy of Sciences and measured for Ca, K, Mg, Na, Si, Ag, Hg, Tl, Al, As, B, Ba, Be, Cd, Co, Cr, Cu, Fe, Mn, Mo, Ni, P, Pb, Sb, Se, Sn, Sr, Ti, V, and Zn by ICP-MS 7900 (Agilent, US), as described in [22].

### 2.4 Measurement of Antibiotic Concentration

Concentrations of amoxicillin and tetracycline were measured using 500 mL of stored filtrate using the Amoxicillin ELISA kit (Shenzhen Lvshiyuan Biotechnology Co., Ltd, China) and Tetracycline ELISA kit (Shenzhen Lvshiyuan Biotechnology Co., Ltd, China), according to the manufacturer’s instructions. The procedures of the two kits were technically the same. Briefly, samples and standard solutions with known antigen (amoxicillin/tetracycline) concentration were separately mixed with corresponding enzyme conjugates (enzyme-labelled antibodies) on a micro-well plate where micro-well strips were pre-coated with coupling antigens. The antigens in the sample and the coupling antigens would compete for the enzyme conjugates. After removing the mixed solution and adding TMB substrate for coloration, the enzyme conjugates bound with sample antigens were removed and only those bound with pre-coated antigens remained, which led to higher optical density (OD) value in the sample with lower antigen concentration. The OD values of the solutions generated from samples and standard solutions were measured with the wavelength of 450nm. All procedures above were performed in duplicate for each sample and standard solution. A standard curve of absorption versus antigen concentration was generated with the OD values from the standard solutions, and the OD values from the samples were compared to the standard curve to get the antigen concentration.

### 2.5 Environmental Risk Assessment of Antibiotics

To assess the potential risk of antibiotics in the environment, a hazard quotient (HQ) was used, which is the ratio between the measured environmental concentration (MEC) of the antibiotic and the predicted no-effect concentration (PNEC) [17]. HQ values >1.0 indicate a potential risk to the aquatic system [23]. This study adopts PNEC values proposed by Bengtsson-Palme and Larsson, 2016 (0.25 μg/L for amoxicillin and 1 μg/L for tetracycline) [24].

### 2.6 Statistical Method

ARG screening data was calculated to relative abundance using the median Cq value of *tetE* in Sample S7 as a reference with the following formula:

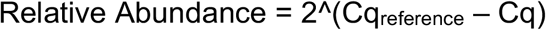

*TetBP, ampr*, and *tetO* qPCR data was calculated to absolute abundance using a standard curve derived by serially diluting (10-fold) synthetic genes at a range of 10^12^ copies/mL to 10^2^ copies/mL.

Concentrations of amoxicillin and tetracycline were calculated using a standard curve derived from standard solutions provided in the commercial kits.

All figures were generated using R (version 4.0 and 4.1). Figure 1(a) used country border data and river data from the rnaturalearth package. Population density data was collected by the Ministry of Civil Affairs of the People’s Republic of China [25] and Statistical Bureau of Republic of China [26]. Figure 1(b) adopted the Stamen map through the ggmap package.

**Figure 1:**
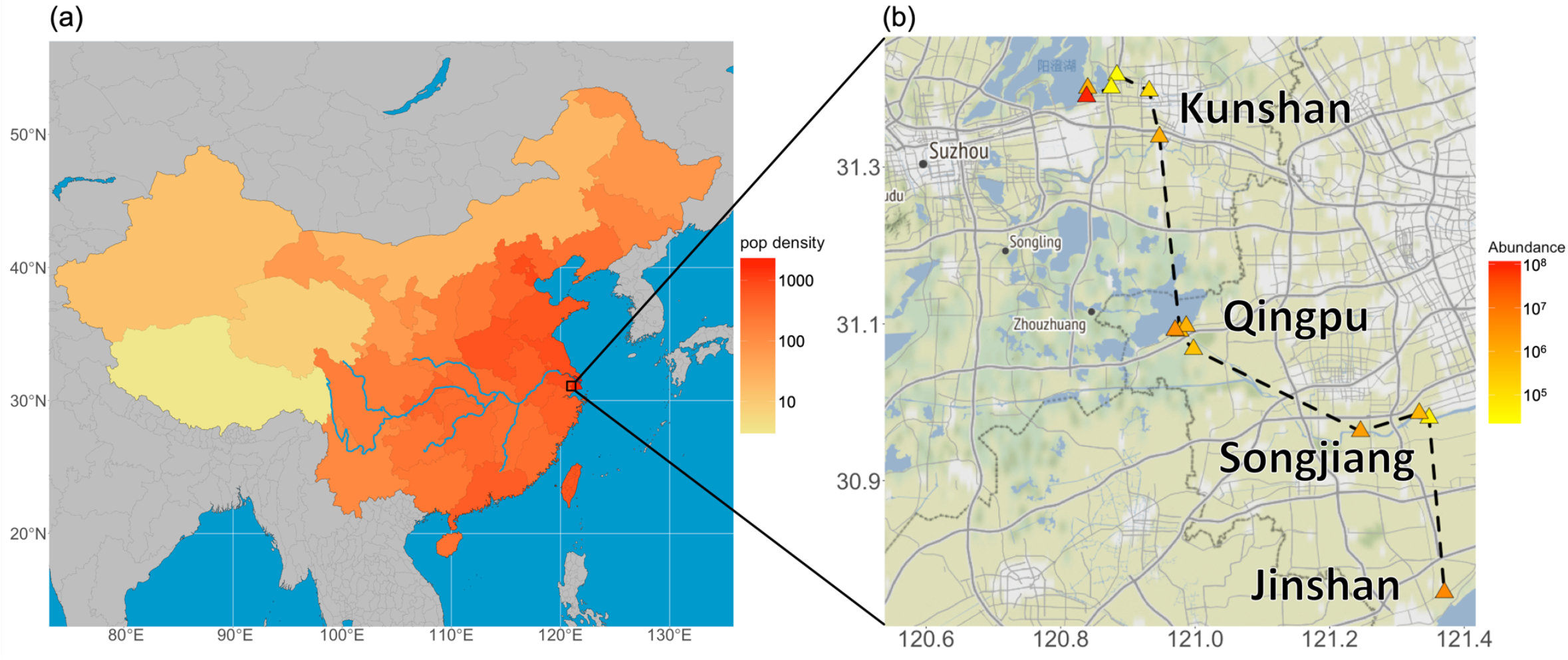
Map of sampling area. (a) Map of China with population density in 2020. Unit of population density: persons/square kilometre. The blue line represents the Yangtze River. (b) Spatial distribution of samples and absolute abundance of total ARGs in each sample. Unit of total ARG abundance: copies/mL. The black dash represents the sampling lane.

Phosphorus, chromium, manganese, calcium, and strontium were used to predict *tetBP, ampr*, and *tetO* abundance through ridge regression. Ridge regression is an algorithm that produces stable estimation of the least-squares coefficients of multiple-regression models with the condition of high collinearity [27]. Because *tetBP* contained an outlier with an abundance several magnitudes higher than the others, *tetBP* was re-scaled through the calculation of log_10_(Abd+1).

## 3. Results

### 3.1 Abundance of Antibiotic Resistance Genes

The spatial distribution of the samples and absolute abundance of total ARG (sum of *tetBP, ampr, tetO*) are shown in Figure 1. Notably, an ARG hotspot (S10) was found in Yangcheng Lake, Kunshan. The hotspot contains an elevated amount of *tetBP* (Figure S1).

The mean absolute abundance and composition of ARGs in the four regions are shown in Figure 2. The ARG *ampr* accounts for the majority of detected concentrations in all the regions. However, all the results shown a large standard deviation, indicating high variability within each region.

**Figure 2:**
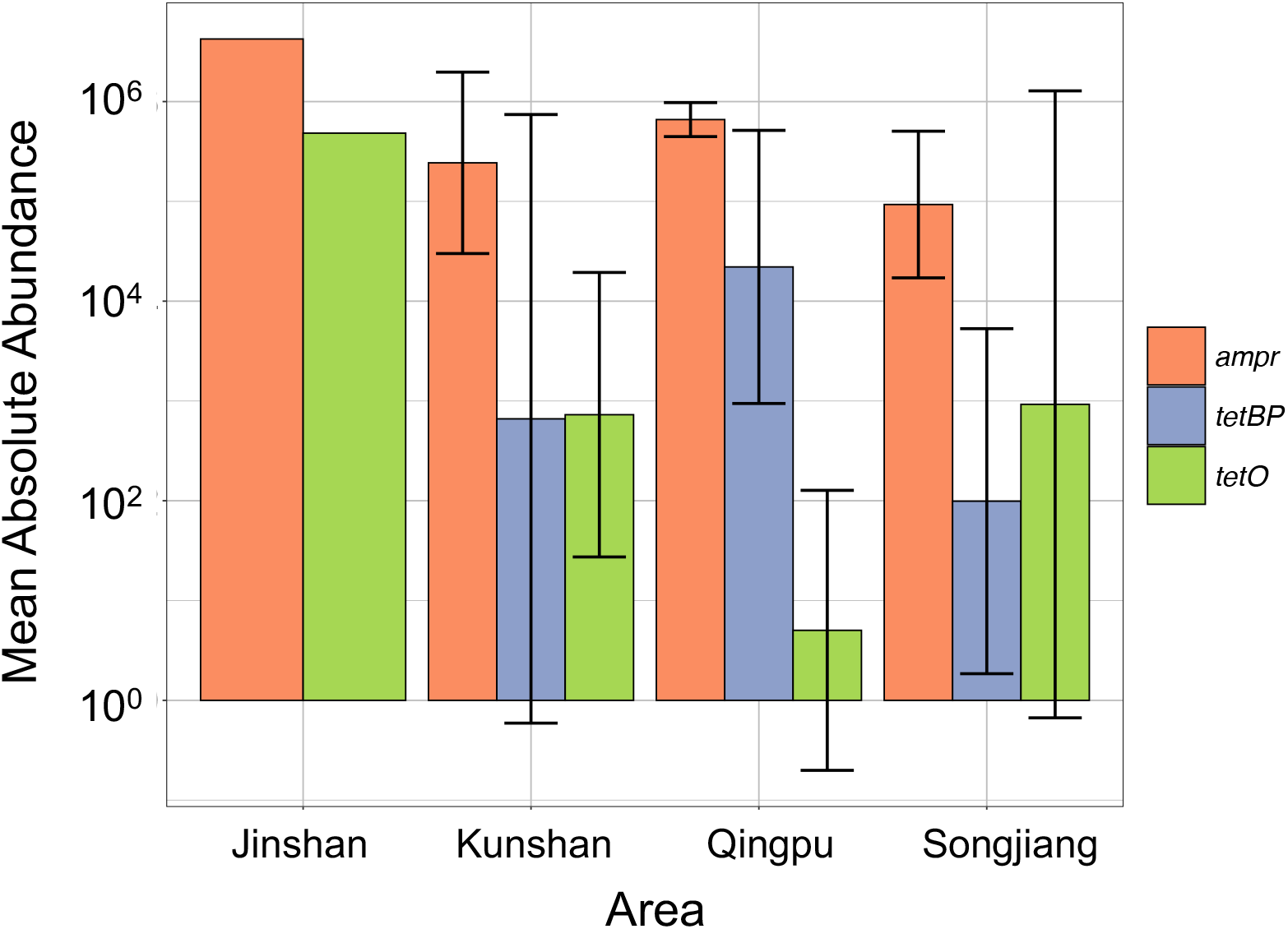
Mean absolute abundance of ARGs in the four sampling regions. Unit of ARG abundance: copies/mL.

### 3.2 Dissolved Elements & Water Quality

In general, all of the samples had a very low heavy metal concentration as shown in Table 1 (according to the Environmental Quality Standard for Surface Water of People’s Republic of China [28]). However, Sample S8 collected from drinking water reserve in Songjiang District, Shanghai had an iron concentration of 417.436 μg/L that exceeds the standard limit of 300 μg/L (Table 2). Moreover, though the concentration of phosphorus in many samples was relatively high (Table 1), the values were still under the standard limits. One exception again was the Songjiang sample (237.872 μg/L) which exceeds the phosphorus limit of 200 μg/L for drinking water (Table 1).

**Table 1:** Water Quality Rank (according to Environmental Quality Standard for Surface Water of PRC [28]) and Concentration (μg/L) of Dissolved Elements. Rank I is applicable for national nature reserves. Rank II is for first-class drinking water reserves. Rank III is for second-class drinking water reserves. Rank IV is for industry water and non-exposure entertainment water sources. Rank V is for agricultural water sources.

| Sample | Area | P |  | Cu |  | Zn |  | As |  | Se |  | Hg |  | Cd |  | Pb |  | Cr |  |
| --- | --- | --- | --- | --- | --- | --- | --- | --- | --- | --- | --- | --- | --- | --- | --- | --- | --- | --- | --- |
| S1 | Qingpu | III | 128.006 | I | 0.174 | I | ND | I | 4.386 | I | ND | I | 0.047 | I | 0.018 | I | 2.922 | I | 0.973 |
| S2 | Qingpu | III | 136.442 | I | 0.427 | I | ND | I | 4.345 | I | ND | I | 0.025 | I | 0.012 | I | 0.791 | I | 1.28 |
| S3 | Qingpu | II | 94.828 | I | 0.433 | I | 7.425 | I | 4.288 | I | ND | I | 0.021 | I | 0.026 | I | 1.372 | I | 0.742 |
| S4 | Qingpu | II | 79.516 | I | 0.056 | I | 1.762 | I | 5.011 | I | ND | I | 0.034 | I | 0.021 | I | 2.112 | I | 0.752 |
| S5 | Kunshan | II | 37.645 | I | ND | I | ND | I | 2.329 | I | ND | I | ND | I | 0.017 | I | 1.914 | I | 0.522 |
| S7 | Jinshan | V | 458.891 | I | ND | I | 5.135 | I | 3.743 | I | ND | I | ND | I | 0.004 | I | 0.668 | I | 2.074 |
| S8* | Songjiang | IV | 237.872 | I | ND | I | 2.272 | I | 3.626 | I | 0.535 | I | ND | I | 0.011 | I | 0.831 | I | 0.6 |
| S9 | Songjiang | III | 180.972 | I | 0.529 | I | 3.414 | I | 3.276 | I | ND | I | ND | I | 0.023 | I | 1.205 | I | 1.33 |
| S10 | Kunshan | II | 34.28 | I | ND | I | ND | I | 2.359 | I | ND | I | ND | I | ND | I | 0.457 | I | ND |
| S11 | Kunshan | II | 30.058 | I | ND | I | ND | I | 2.442 | I | ND | I | ND | I | 0.005 | I | 0.642 | I | ND |
| S12* | Kunshan | II | 45.482 | I | ND | I | 9.226 | I | 2.708 | I | ND | I | ND | I | ND | I | 0.332 | I | 0.264 |
| S13* | Kunshan | II | 24.372 | I | ND | I | ND | I | 2.217 | I | ND | I | ND | I | 0.004 | I | 0.646 | I | 0.113 |
| S14 | Kunshan | IV | 258.324 | I | 0.275 | I | 0.72 | I | 2.712 | I | ND | I | ND | I | 0.022 | I | 0.967 | I | 0.351 |
| S15 | Kunshan | III | 114.272 | I | 2.07 | I | 11.143 | I | 2.309 | I | ND | I | ND | I | 0.031 | I | 2.364 | I | 1.125 |

\*Labelled samples are from drinking water reserves. ND: Not detected.

**Table 2:** Additional requirements from Environmental Quality Standard for Surface Water of PRC for drinking water reserves.

| Sample | Area | Mo |  | Co |  | Be |  | B |  | Sb |  | Ni |  | Ba |  | Ti |  | V |  | Tl |  | Fe |  |
| --- | --- | --- | --- | --- | --- | --- | --- | --- | --- | --- | --- | --- | --- | --- | --- | --- | --- | --- | --- | --- | --- | --- | --- |
| S8 | Songjiang | W | 3.538 | W | 0.326 | W | ND | W | 99.268 | W | 0.751 | W | 0.548 | W | ND | W | 6.237 | W | 2.632 | W | ND | E | 417.436 |
| S12 | Kunshan | W | 3.302 | W | 0.153 | W | ND | W | 119.726 | W | 1.407 | W | 1.998 | W | ND | W | 1.985 | W | 2.021 | W | ND | W | 153.924 |
| S13 | Kunshan | W | 2.453 | W | 0.075 | W | ND | W | 115.066 | W | 0.742 | W | 1.312 | W | ND | W | 0.596 | W | 0.856 | W | ND | W | 61.913 |

“W” means concentration is within the limit for drinking water reserves, and “E” means exceeding the requirement; ND: Not detected.

### 3.3 Measurement of Antibiotic Concentration and Risk Assessment

Antibiotic concentrations are shown in Table 3. Unexpectedly, except for the sample from Jinshan contains 0.13 μg/L of tetracycline, antibiotics were not detected in any other samples. Also, the HQ value tetracycline concentration in Jinshan is 0.13, indicating no risk.

**Table 3:** Concentrations of amoxicillin and tetracycline (unit: μg/L).

| Sample | Area | Amoxicillin | Tetracycline |
| --- | --- | --- | --- |
| S1 | Qingpu | ND | ND |
| S2 | Qingpu | ND | ND |
| S3 | Qingpu | ND | ND |
| S4 | Qingpu | ND | ND |
| S5 | Kunshan | ND | ND |
| S6 | Songjiang | ND | ND |
| S7 | Jinshan | ND | ND |
| S8 | Songjiang | ND | ND |
| S9 | Songjiang | ND | ND |
| S10 | Kunshan | ND | ND |
| S11 | Kunshan | ND | ND |
| S12 | Kunshan | ND | ND |
| S13 | Kunshan | ND | ND |
| S14 | Kunshan | ND | ND |
| S15 | Kunshan | ND | ND |

ND: Not detected. Detection limit: 1 µg/L.

### 3.4 Correlation of ARGs and Environmental Risk Factors

Figure 3 shows the correlation of ARGs with salinity, temperature, and dissolved elements. Water temperature and salinity show weak to little correlation with ARGs. pH appears to have a weak negative correlation with *tetO* and *ampr* (r = -0.58, - 0.55). Interestingly, phosphorus, chromium, and manganese exhibit a strong positive correlation with *tetO* (r = 0.79, 0.64, 0.63). Calcium has a strong negative correlation with *ampr* (r = -0.70). And strontium has strong negative correlation with both *tetO* and *ampr* (r = -0.71, -0.70). Linear regression of *tetO* and *ampr* with phosphorus, chromium, manganese, calcium, and strontium is shown in Figure 4. These five elements explain 71.2% variability of *ampr*, 79.1% variability of *tetO*, and 36.0% variability of log-scaled *tetBP*, in a ridge regression model.

**Figure 3:**
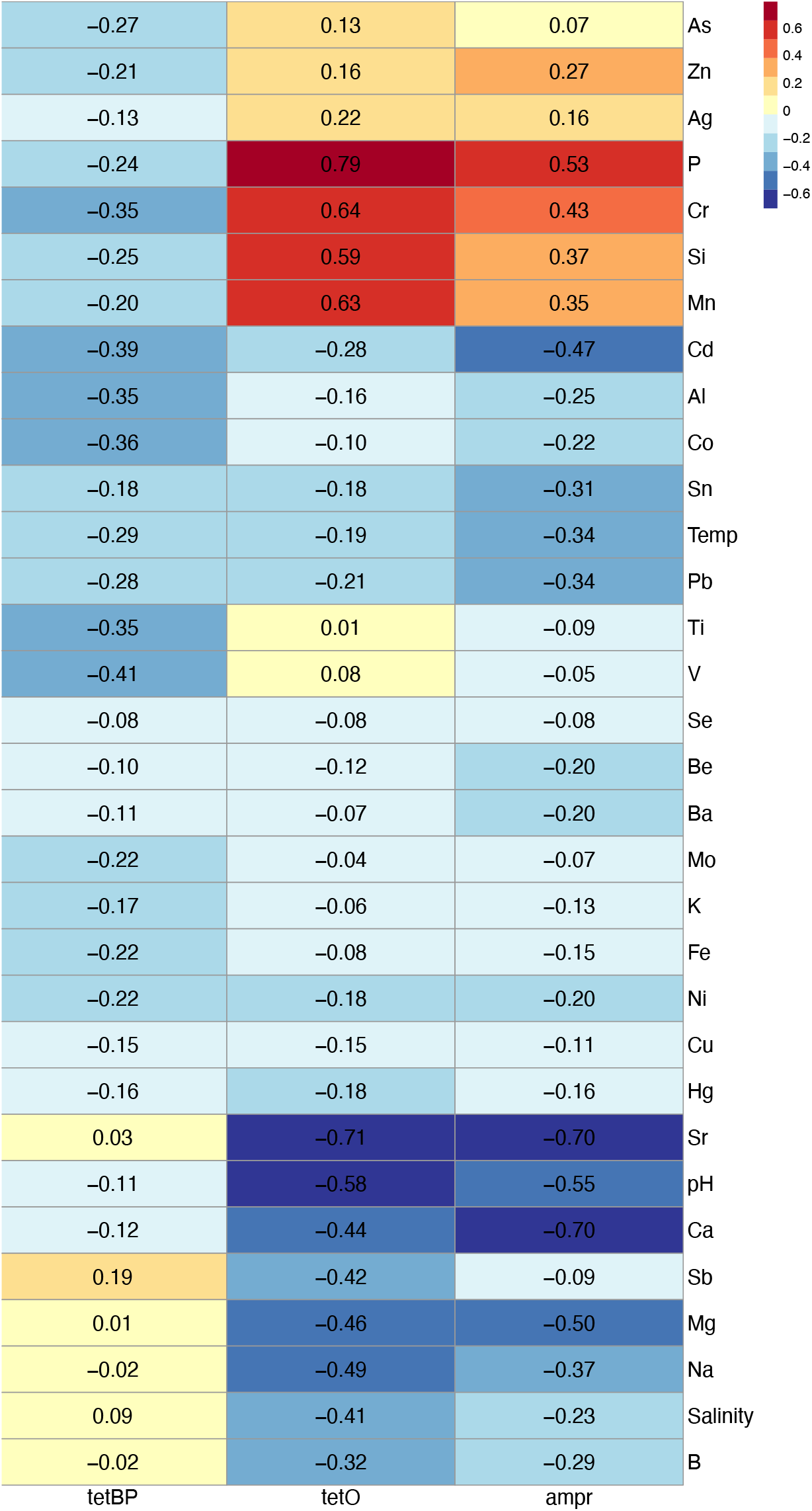
Heatmap of correlation of ARG concentrations and environmental parameters.

**Figure 4:**
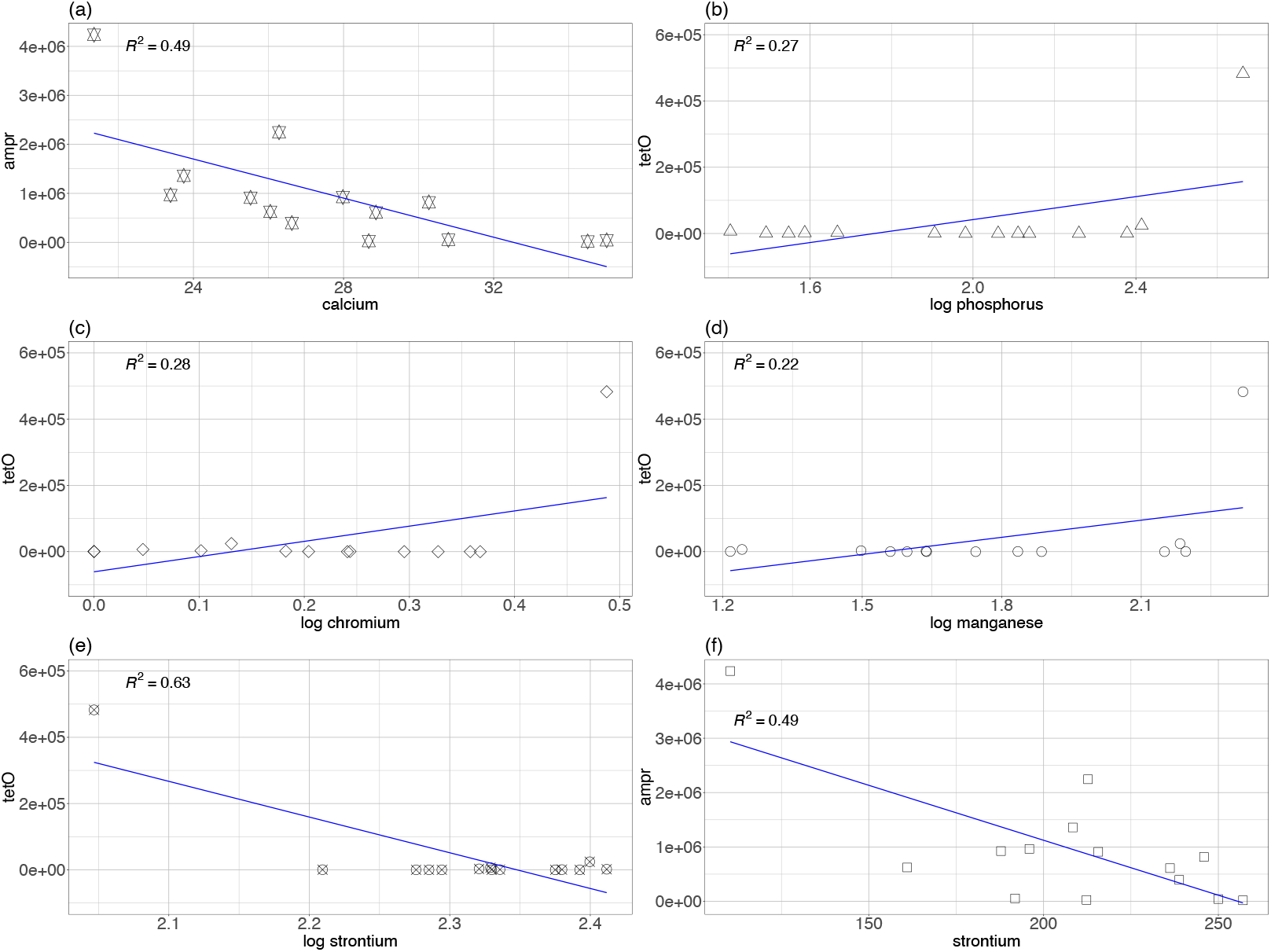
Linear regression of dissolved elements with ARGs (a) Calcium and *ampr*. (b) Phosphorus and *tetO*. (c) Chromium and *tetO*. (d) Manganese and *tetO*. Unit: tetO (copies/mL). Unit: ampr (copies/mL), tetO (copies/mL), calcium (mg/L), phosphorus (μg/L), chromium (μg/L), manganese (μg/L), strontium (μg/L).

## 4. Discussion

This study aims to understand what environmental factors might contribute to the prevalence of ARGs in main water bodies in the Yangtze Delta. In summary, a high abundance of ARGs was detected and several dissolved elements (phosphorus, chromium, manganese, calcium, and strontium) were potentially key contributors of ARGs.

### 4.0 Interpretation of the abundance of ARGs

As shown in Figure 1 and Figure 2, a high abundance of ARGs was detected. Compared with historical data of Huangpu River collected in July 2013 [29], the ARGs abundance in Huangpu River detected in this study was several magnitudes higher. Specifically, historical data shows that the abundance of tetracycline-resistant genes was at the magnitude of 10 to 10^4^ copies/mL [29], but in this study *tetO* abundance in Huangpu River reached up to 10^6^ copies/mL (Figure 2). The hotspot sample from Yangcheng Lake, Kunshan, contained *tetBP* at the magnitude of 10^8^ copies/mL (Figure 1). This finding suggests there could have been a surge of antibiotic resistant bacteria in the Yangtze Delta during the past decade.

What is more, the hotspot and large standard deviations of ARG concentrations imply that there might be point sources of ARG pollution, which elevated ARG concentrations locally (e.g., hotspot of Sample S10), but could not disperse over a wide area.

### 4.1 ARGs and Antibiotics

In contrast with the detection of ARGs, antibiotic measurement results indicate that antibiotic persistence in the environment was limited, with ELISA results showing that all samples contained less than 1 μg/L of amoxicillin and tetracycline (Table 3). This finding is consistent with other studies that show antibiotic concentrations in surface water in Shanghai is at the magnitude of ng/L [13-15, 29-31]. This indicates, despite low antibiotic risk, ARGs remain persistent in the aquatic system.

Notably, persistence of tetracycline resistance was also reported in coastal regions of Southeast China, which is geologically close to this study’s sampling region [32]. Though other mechanisms may also contribute to persistence of tetracycline resistance (e.g., compensatory evolution), existing evidence supports co-selection between resistance genes is the most likely cause of persistence of tetracycline resistance [6], which will be discussed in following parts.

Additionally, it is worth noting that overall antibiotic levels in the Yangtze Delta were observed to vary in accordance to seasons, with the number of detected antibiotics and their detection frequencies being significantly lower in summer than in winter [14, 15, 33, 34], except for tetracyclines [34]. The high flow conditions in summer might dilute the concentration of antibiotics in the surface water [35], and faster degradation of antibiotics in summer could also contribute to the low concentration of antibiotics detected [34, 36]. The results of this study do not guarantee a risk of antibiotics during the winter.

Though the correlation coefficient of ARGs and antibiotics cannot be calculated, considering the low level of antibiotic concentrations, it might be more prudent to focus on direct remediation strategies for removing ARGs, instead of focusing only on decreasing antibiotic contamination in surface water of the Yangtze Delta.

### 4.2 ARGs and Nonmetal Elements

As shown in Figure 3 and Figure 4, phosphorus exhibits a strong positive correlation with *tetO*. One possible explanation for this correlation is that phosphorus, as an essential biological element, enriches bacterial growth in freshwater [37, 38]. The abundance of ARGs proliferate as the quantity of bacterial cells increases. This positive connection between phosphorus and ARGs is also reported by other studies in eutrophic environment [39, 40]. Though additional assessment is needed to better understand the relationship between phosphorous concentrations and ARG, the findings seem to suggest that regulating phosphorous eutrophication could be a practical approach to manage ARG development in surface water. One similar proposal is also given by Wang et al. 2020 [40].

### 4.3 ARGs and Heavy Metals

As shown in Figure 3 and Figure 4, chromium and manganese exhibit a strong positive correlation with *tetO*. The positive correlation of chromium could be a result of co-selection between tetracycline-resistant genes and chromium-resistant genes though co-resistance mechanism. Co-resistance describes the spatial association of HMRGs and ARGs when they are located on the same genetic elements, such as plasmids and transposons. This association means either heavy metals or antibiotics can be the selection pressure for both resistance genes. A previous study cured plasmids from *Salmonella abortus equi* strains and found the plasmids were resistant to ampicillin, arsenic, chromium, cadmium and mercury simultaneously [41].

However, co-selection of manganese and ARGs is rarely reported. Further research is needed to confirm whether manganese truly has co-selective pressure on ARGs.

### 4.4 ARGs and Light Metals

A negative correlation was found between *ampr* and calcium, and strontium had a strong negative correlation with both *tetO* and *ampr* (Figure 3 & Figure 4).Interestingly, as noted, a previous study reported magnesium (Mg) could suppress resistant bacterial community [8]. Magnesium, calcium, and strontium all belong to Group 2A on the periodic table. This means the three elements have similar chemical properties, so it is possible that the three elements have a potential inhibition effect on ARG through similar mechanisms.

Although negative correlation of strontium may be explained by its cytotoxicity on bacteria [42, 43], molecular biology studies have reported selective inhibition effects of magnesium and calcium on antibiotic-resistant bacteria. Magnesium and calcium may involve directly in the destabilization of the *Staphylococcus aureus* cell membrane [44]. Calcium and magnesium can form a complex with Cardiolipin (CL), one major component of the bacterial cell membrane, thus destabilizing the cell membrane and eventually causing cell death [44]. In addition, calcium could suppress bacterial growth through inhibiting peptidoglycan-layer (PG-layer) synthesis or destabilizing the cell membrane. PG-layer synthesis involves transpeptidase (TP) enzymes, including D,D-TP and L,D-TP. These TP enzymes are the target of many antibiotic β-lactams [45]. Calcium hinders the binding of L,D-TP to PG-stem by inducing the formation of L,D-TP dimer [45].

This finding reveals the possibility of developing Group 2A light metals as reagents to control ABR. For example, as one of the major elements in nature, calcium salts are of low toxicity and eco-friendly. Calcium-based reagent might be capable of suppressing antibiotic resistant bacteria in natural environment if added to natural surface water (e.g., exposure recreational water).

### 4.5 Limitations

Considering the reported seasonal variance in antibiotic concentrations [14, 15, 33, 34], the findings on antibiotic concentrations can only be generalizable to surface water of the Yangtze Delta in summer. Also, impacted by the outbreak of COVID-19 Delta strain in China in 2021 Summer, the sampling stage of this study was emergently terminated, ahead of schedule. The unexpected termination led to a small sample size (15 samples). The small sample size decreases statistical power and increases potential error in the results. Finally, ARG concentrations were of large standard deviations in all four subregions. Though this could be explained as indication of point sources of ARG pollution, one alternative explanation is there was degradation of DNA during the experiments.

## 5. Conclusion

This study detected a high abundance of ARGs in surface water samples collected from the Yangtze Delta, despite corresponding low antibiotic and low heavy metal concentration. Phosphorus, chromium, manganese, calcium, and strontium were identified as potential key contributors of ARGs in the Yangtze Delta. Reduction of ARG abundance could be realized through decreasing the concentration of phosphorus in surface water. In addition, Group 2A light metals (e.g., magnesium and calcium) were identified to have a potential ARG suppression effect, though future studies are still needed to further evaluate this association.

## Data Availability

All data produced in the present work are contained in Supporting Data with this preprint.

## 6. Acknowledgements

The authors thank Dr. Huansheng Cao for providing the positive controls for the quantitative PCR experiments and thank Dr. Yingxue Li for her help in measuring the antibiotic concentrations.

This work was funded by Interdisciplinary Health Research Fund at the Global Health Research Center, Duke Kunshan University, Kunshan, China.

