## Supplementary Table and Figures for "Assessment of Environmental Factors Associated with Antibiotic Resistance Genes (ARGs) in the Yangtze Delta, China"

**Benjamin D. Anderson, MPH, PhD, CPH**

Assistant Professor

Department of Environmental and Global Health, College of Public Health and Health Professions, and Emerging Pathogens Institute, University of Florida, Gainesville, Florida, 32610 United States

**Shixin Xu, PhD**

Assistant Professor of Mathematics

Division of Natural and Applied Science, Duke Kunshan University, Kunshan, Jiangsu, 215300, China

Global Health Research Center, Duke Kunshan University, Kunshan, Jiangsu, 215300, China

**Table and Figure Legend**

Table S1: Sequences and cycling condition of quantitative PCR experiments of ARG primers.

Figure S1: Relative abundance of 22 ARGs detected in screening step. (a) The sum of relative abundance of ARGs. (b) The prevalence of ARGs in the samples. Negative results are represented as grey blocks.

Figure S2: Correlation plot of phosphorus, chromium, manganese, calcium, and strontium.

| | Orientation | Sequence | Length | Annealing Temperature | References |
|---|---|---|---|---|---|
| *tetq* | F | AGAATCTGCTGTTTGCCAGTG | 169bp | 55 | [1] |
| | R | CGGAGTGTCAATGATATTGCA | | | |
| *sul3* | F | TCCGTTCAGCGAATTGGTGCAG | 128bp | 60 | [2] |
| | R | TTCGTTCACGCCTTACACCAGC | | | |
| *ermB* | F | GGCATTTAACGACGAAACTGGC | 236bp | 57 | [2] |
| | R | CGCATGGCTTTCAAAAACCAC | | | |
| *ampr* | F | GAGTTTTCGTTCCACTGAGCGTC | 274bp | 60 | [3] |
| | R | TTAGCAGAGCGAGGTATGTAGGCG | | | |
| *tetA* | F | GCTACATCCTGCTTGCCTTC | 210bp | 55 | [4] |
| | R | CATAGATCGCCGTGAAGAGG | | | |
| *tetC* | F | CTTGAGAGCCTTCAACCCAG | 418bp | 55 | [4] |
| | R | ATGGTCGTCATCTACCTGCC | | | |
| *tetE* | F | GTTATTACGGGAGTTTGTTGG | 278bp | 55 | [4] |
| | R | AATACAACACCCACACTACGC | | | |
| *tetG* | F | GCTCGGTGGTATCTCTGCTC | 468bp | 55 | [4] |
| | R | AGCAACAGAATCGGGAACAC | | | |
| *tetK* | F | CGAAAACAGACTCGCCAATC | 169bp | 55 | [4] |
| | R | TCCATAATGAGGTGGGGC | | | |
| *tetL* | F | TCGTTAGCGTGCTGTCATTC | 267bp | 55 | [4] |
| | R | GTATCCCACCAATGTAGCCG | | | |

| | | | | | |
|---|---|---|---|---|---|
| *tetAP* | F | CTTGGATTGCGGAAGAAGAG | 676bp | 55 | [4] |
| | R | ATATGCCCATTTAACCACGC | | | |
| *tetS* | F | CATAGACAAGCCGTTGACC | 667bp | 55 | [4] |
| | R | ATGTTTTTGGAACGCCAGAG | | | |
| *tetM* | F | ACAGAAAGCTTATTATATAAC | 171bp | 45 | [5] |
| | R | TGGCGTGTCTATGATGTTCAC | | | |
| *tetO* | F | ACGGARAGTTTATTGTATACC | 171bp | 45 | [5] |
| | R | TGGCGTATCTATAATGTTGAC | | | |
| *tetT* | F | AAGGTTTATTATATAAAAGTG | 169bp | 40 | [5] |
| | R | AGGTGTATCTATGATATTTAC | | | |
| *tetW* | F | GAGAGCCTGCTATATGCCAGC | 168bp | 60 | [5] |
| | R | GGGCGTATCCACAATGTTAAC | | | |
| *tetBP* | F | AAAACTTATTATATTATAGTG | 169bp | 40 | [5] |
| | R | TGGAGTATCAATAATATTCAC | | | |
| *sulI* | F | CGCACCGGAAACATCGCTGCAC | 163bp | 56 | [6] |
| | R | TGAAGTTCCGCCGCAAGGCTCG | | | |
| *sulII* | F | TCCGGTGGAGGCCGGTATATGG | 191bp | 61 | [6] |
| | R | CGGGAATGCCATCTGCCTTGAG | | | |
| *ereA* | F | AACACCCTGAACCCAAGGGACG | 420bp | 52 | [7] |
| | R | CTTCACATCCGGATTCGCTCG | | | |
| *ereB* | F | AGAAATGGAGGTTCATACTTACCA | 546bp | 52 | [7] |

| | | | | | |
|---|---|---|---|---|---|
| | R | CATATAATCATCACCAATGGCA | | | |
| *mphA* | F | AACTGTACGCACTTGC | 837bp | 52 | [7] |
| | R | GGTACTCTTCGTTACC | | | |
| $bla_{CTX\_M}$ | F | ATGTGCAGYACCAGTAARGT | 593bp | 50 | [8] |
| | R | TGGGTRAARTARGTSACCAGA | | | |
| $bla_{TEM}$ | F | KACAATAACCCTGRTAAATGC | 936bp | 58 | [8] |
| | R | AGTATATATGAGTAAACTTGG | | | |
| $bla_{SHV}$ | F | TTTATCGGCCYTCACTCAAGG | 930bp | 58 | [8] |
| | R | GCTGCGGGCCGGATAACG | | | |
| $bla_{ampC}$ | F | CCCCGCTTATAGAGCAACAA | 634bp | 58 | [8] |
| | R | TCAATGGTCGACTTCACACC | | | |

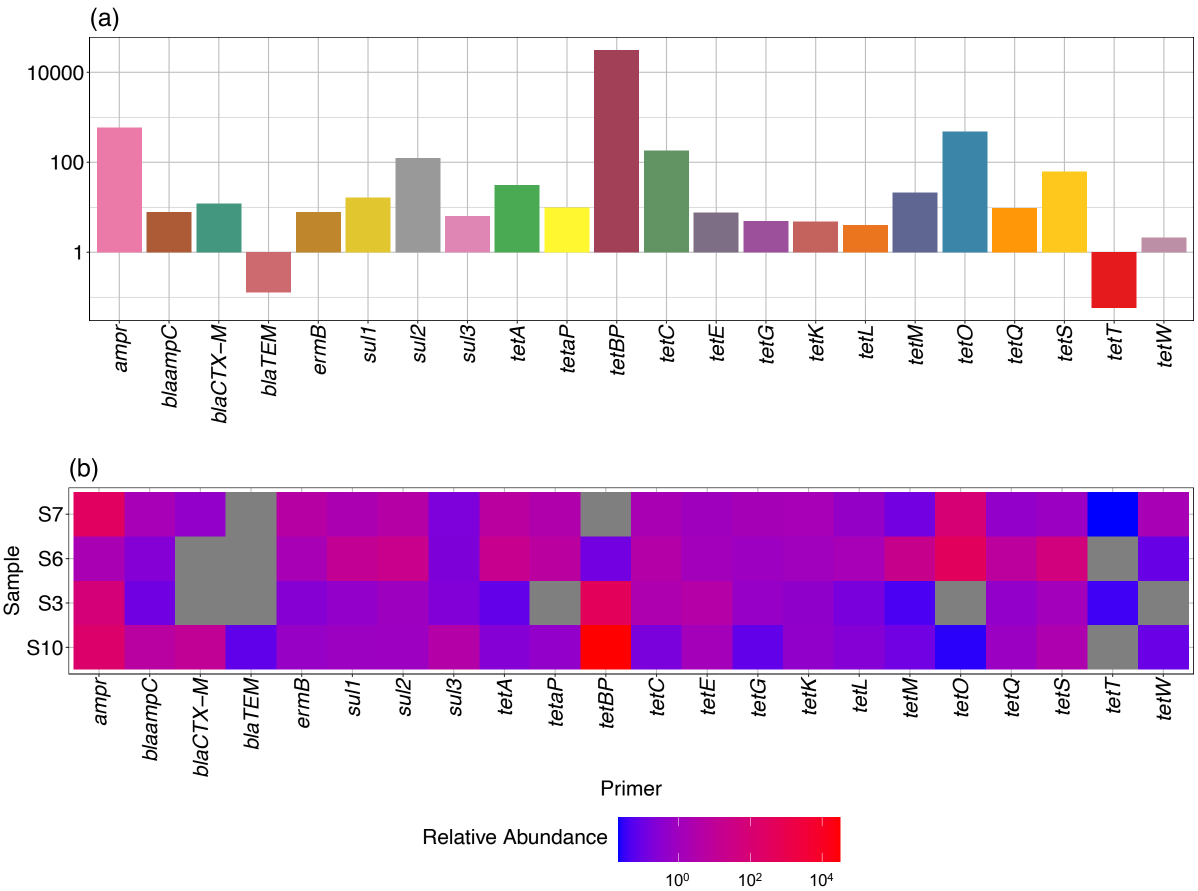
(a)
10000
100
1
ampr
blaampC
blaCTX–M
blaTEM
ermB
sul1
sul2
sul3
tetA
tetaP
tetBP
tetC
tetE
tetG
tetK
tetL
tetM
tetO
tetQ
tetS
tetT
tetW
(b)
Sample
S7
S6
S3
S10
Primer
Relative Abundance
10⁰
10²
10⁴

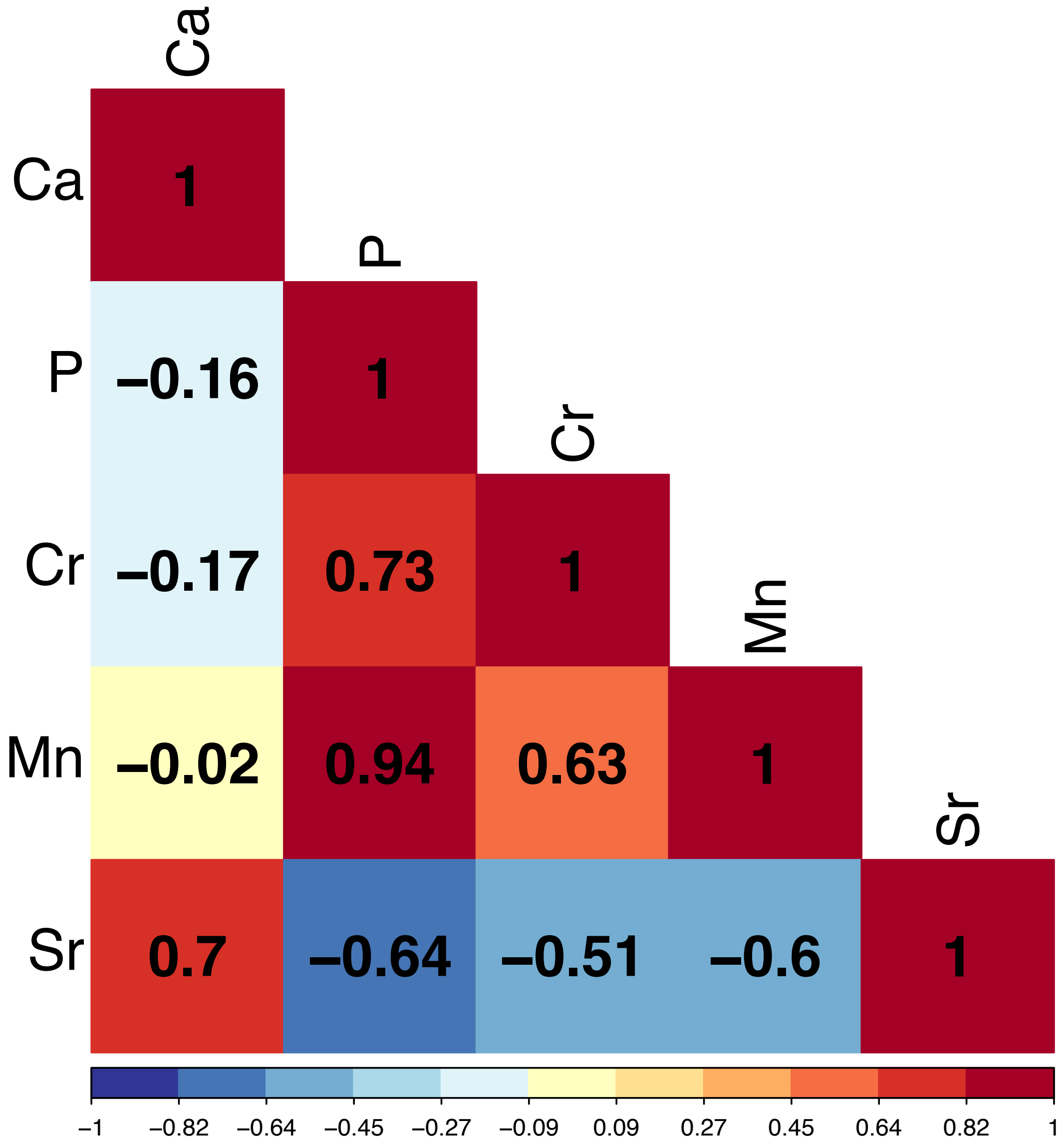

Ca
P
Cr
Mn
Sr
Ca
P
Cr
Mn
Sr
1
−0.16
1
−0.17
0.73
1
−0.02
0.94
0.63
1
0.7
−0.64
−0.51
−0.6
1
−1
−0.82
−0.64
−0.45
−0.27
−0.09
0.09
0.27
0.45
0.64
0.82
1
